# Nosocomial outbreak of SARS-CoV-2 in a “non-COVID-19” hospital ward: virus genome sequencing as a key tool to understand cryptic transmission

**DOI:** 10.1101/2021.02.20.20248421

**Authors:** Vítor Borges, Joana Isidro, Filipe Macedo, José Neves, Luís Silva, Mário Paiva, José Barata, Judite Catarino, Liliana Ciobanu, Sílvia Duarte, Luís Vieira, Raquel Guiomar, João Paulo Gomes

## Abstract

**Background:** Dissemination of severe acute respiratory syndrome coronavirus 2 (SARS-CoV-2) in healthcare institutions affects both patients and health-care workers (HCW), as well as the institutional capacity to provide essential health services.

**Methods:** We conducted an investigation of a cluster of SARS-CoV-2 positive cases detected in a “non-COVID-19” hospital ward during Summer 2020. The magnitude of the nosocomial outbreak was disclosed by massive testing, challenging the retrospective reconstruction of the introduction and transmission events. An in-depth contact tracing investigation was carried out to identify the contacts network during the 15-day period before the screening. In parallel, positive SARS-CoV-2 RNA samples were subjected to virus genome sequencing.

**Results:** Of the 245 tested individuals, 48 (21 patients and 27 HCWs) tested positive for SARS-CoV-2. HCWs were mostly asymptomatic, but the mortality among the vulnerable patient group reached 57.1% (12/21). Phylogenetic reconstruction revealed that all cases were part of the same transmission chain, thus confirming a single origin behind this nosocomial outbreak. By combining vast epidemiological and genomic data, including analysis of emerging minor variants, we unveiled a scenario of silent SARS-CoV-2 dissemination within the hospital ward, mostly driven by the close contact within the HCWs group and between HCWs and patients. This investigation triggered enhanced prevention and control measures, leading to a more timely detection and containment of novel nosocomial outbreaks.

**Conclusions:** The present study shows the benefit of combining genomic and epidemiological data for the investigation of complex nosocomial outbreaks, and provides valuable data to minimize the risk of transmission of COVID-19 in healthcare facilities.

**Short summary:** SARS-CoV-2 nosocomial outbreaks are of utmost public health concern. Here, we performed an in-depth investigation of a high-fatality rate nosocomial outbreak by combining vast genomic and epidemiological data, providing valuable information to understand cryptic transmission of SARS-CoV-2 within healthcare institutions.

## INTRODUCTION

The pandemic coronavirus SARS-CoV-2 (Severe Acute Respiratory Syndrome Coronavirus 2), the causative agent of COVID-19 [1, 2], is a highly contagious respiratory virus that has rapidly spread globally, having caused more than 105 million reported cases and 2.3 million deaths, as of 13 February 2020 [3]. The clinical presentation of COVID-19 ranges from asymptomatic or mild symptomatic infections to severe respiratory symptoms, multiorgan failure or death, which is more frequent in elderly generally presenting comorbidities [1, 2, 4]. SARS-CoV-2 transmission within healthcare institutions is of utmost public health concern at global level [5-8], with several studies reporting high infection rates among healthcare workers (HCW) and/or hospitalized patients [9-13]. Nosocomial COVID-19 outbreaks can have a tremendous negative impact at individual health level, such as detrimental effects on HCW well-being and increased morbidity and mortality among patients. Also, at community level, they implicate the loss of institutional capacity, namely staff and resources, to provide non-COVID-19 and COVID-19 clinical assistance to the community. In this context, several recommendations have been launched by national and international health authorities [5, 6] to place robust preventive (e.g., guarantee sufficient supplies of personal protective equipment; promote routine testing among HCW) and control measures to timely detect and limit SARS-CoV-2 transmission within healthcare institutions. With the recent advances in sequencing technologies, it is now possible to strengthen outbreak investigations by performing timely virus genome sequencing [10, 14-16]. This tool can be key not only to detect and reconstruct nosocomial transmission chains, but also to guide outbreak control actions and/or prospective preventive measures. Here, we performed an in-depth investigation of a high-fatality rate nosocomial outbreak through the combination of vast genomic and epidemiological data to unveil routes and modes of transmission of SARS-CoV-2 within a non-COVID-19 hospital ward.

## METHODS

### Study population, specimen collection and RT-PCR testing

This study describes a COVID-19 outbreak occurring in a large hospital (313 beds) from the Regional Health Administration of the Lisbon and Tagus Valley (Portugal) that employs around 1500 employees and provides healthcare assistance to around 250000 people. The outbreak occurred in a hospital ward (36 rooms, 73 beds) dedicated to internal medicine (“non-COVID-19” ward B) in Summer 2020. The epidemiological investigation covered 348 health-care workers and 92 patients. SARS-CoV-2 laboratory tests were performed on a total of 245 individuals following the international recommendations. Briefly, naso and oropharyngeal swabs were collected from patients and HCW, and RNA was subsequently extracted using m2000sp Abbott and then subjected to real-time reverse transcription PCR (RT-PCR) assay targeting using m2000rt Abbott. SARS-CoV-2 positive RNA samples were sent to the National Institute of Health (INSA) Dr. Ricardo Jorge for SARS-CoV-2 whole-genome sequencing and bioinformatics analysis.

### Genome sequencing and bioinformatics

Genome sequencing was performed at the INSA following an amplicon-based whole-genome amplification strategy using tiled, multiplexed primers, according to the ARTIC network protocol (https://artic.network/ncov-2019; https://www.protocols.io/view/ncov-2019-sequencing-protocol-bbmuik6w) [17]. In brief, after cDNA synthesis, whole-genome amplification was performed using two separate pools of tiling primers [pools 1 and 2; primers version V3 (218 primers) was used for all samples: https://github.com/artic-network/artic-ncov2019/tree/master/primer_schemes/nCoV-2019]. The two pools of multiplexed amplicons were then pooled for each sample, followed by post PCR clean up and Nextera XT dual-indexed library preparation, according to the manufacturers’ instructions. Sequencing libraries were paired-end sequenced (2×150bp) on an Illumina NextSeq 550 apparatus, as previously described [18]. All bioinformatics analysis (from reads quality control to variant detection/inspection, sequence consensus generation and minor variants analysis) was conducted using the online platform INSaFLU (https://insaflu.insa.pt/) [19], as previously described [18]. The obtained mean depth of coverage was 4248-fold (ranging between 1001- and 5838-fold). All outbreak-associated genome sequences included in the study had >88% of the genome covered by at least 10-fold. Regions with depth of coverage below this threshold were automatically masked in INSaFLU pipeline by placing undefined bases “N” in the consensus sequence (whenever these regions enroll SNP phylogenetic markers, these were inspected to ensure the correct phylogenetic placement of all genomes). Phylogenetic analysis was performed using the SARS-CoV-2 Nextstrain pipeline [20] version from March 23, 2020 (https://github.com/nextstrain/ncov), as previously described [18]. Coronapp (http://giorgilab.dyndns.org/coronapp/) [21] was applied to refine the impact of mutations at protein level. Clade and lineage assignments were performed using Nextclade (https://clades.nextstrain.org/) and Phylogenetic Assignment of Named Global Outbreak Lineages (Pangolin) (https://pangolin.cog-uk.io/) [22], respectively. Outbreak-related SARS-CoV-2 genome sequences generated in this study were uploaded to GISAID database (https://www.gisaid.org/). Accession numbers can be found in Supplementary Table S1.

## RESULTS AND DISCUSSION

### Detection of a COVID-19 nosocomial outbreak

A patient attended to the emergency department of a large hospital in Portugal on Summer 2020 three days later after being discharged from the same hospital, testing positive for SARS-CoV-2 on re-admission. During the previous hospital stay (in ward A), this patient (patient 35) had had close contact with an inpatient (patient 3) from a “non-COVID-19” internal medicine ward (ward B) in a hemodialysis unit (Figure 1). Patient 3 and a clinician (HCW 36, working in ward B) presenting suggestive symptoms for COVID-19 were screened on the following day, also testing positive. These cases triggered a large screening of patients and HCWs in both wards (Figure 1). Further testing was performed during the following days for individuals having either close contacts with positive cases or compatible symptoms. In total, no cases were detected in ward A among 51 tested individuals, whereas 27 out of 102 HCW and 21 out of 92 inpatients linked to ward B tested positive (Figure 1, Table 1). Most HCWs (all except HCWs 13, 31 and 36) were asymptomatic at the time of testing and all recovered from the infection (Table 1). Although most patients (n=15) were also asymptomatic at the time of testing, the mortality rate among COVID-19 patients was 57.1% (12 / 21) (Table 1).

**Table 1.** **Descriptive characteristics of outbreak-related health-care workers (n = 27) and patients (n = 21) testing positive for SARS-CoV-2 RNA in a large hospital in Portugal, Summer, 2020.**

| **Health-care workers (n= 27)** |  |  |
| --- | --- | --- |
| **Sex** |  |  |
|  | **Male** | **4 (14,3%)** |
|  | **Female** | **24 (85,7%)** |
| **Age, years** |  | **32 (22-54)** |
| **Activity** |  |  |
|  | **Medical** | **3 (11,1%)** |
|  | **Nurse** | **10 (37,0%)** |
|  | **Operational assistant** | **12 (44,4%)** |
|  | **Other** | **2 (7,4%)** |
| **Symptoms\***<br>**(time of testing)** |  |  |
|  | **Yes** | **3 (11,1%)** |
|  | **No** | **24 (88,9%)** |
| **Clinical outcome** |  |  |
|  | **Recovery** | **27 (100%)** |
|  | **Death** | **0 (0%)** |
| **Patients (n= 21)** |  |  |
| **Sex** |  |  |
|  | **Male** | **10 (47,6%)** |
|  | **Female** | **11 (52,4%)** |
| **Age, years** |  | **82 (46-92)** |
|  | **>70** | **19 (90,5%)** |
| **Comorbidities** |  |  |
|  | **Yes** | **18 (85,7%)** |
|  | **No** | **3 (14,3%)** |
| **Symptoms\***<br>**(time of testing)** |  |  |
|  | **Yes** | **6 (28,6%)** |
|  | **No** | **15 (71,4%)** |
| **Clinical outcome** |  |  |
|  | **Recovery** | **9 (42,9%)** |
|  | **Death** | **12 (57,1%)** |
| Data are presented as: n (%) or median (range).<br>Symptoms: Fever, cough or shortness of breath. |  |  |

**Figure 1.**
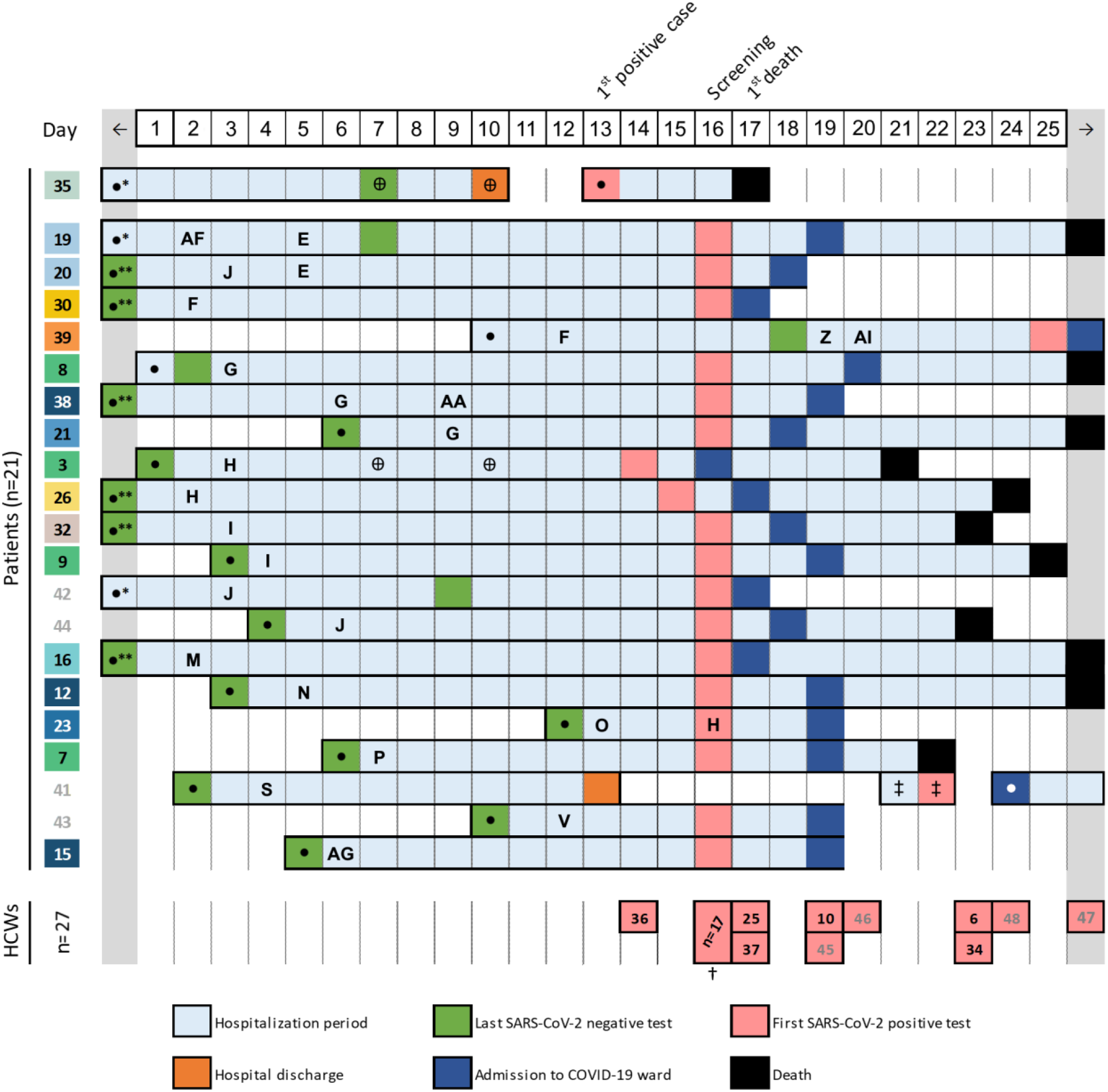
Timeline of events during the SARS-CoV-2 nosocomial outbreak. The figure summarizes the main events occurring during the study period, in which day 1 corresponds to the first day of the 15-day period before the large screening. It depicts the day of hospital admission (•), rooms transfer, sample collection for last negative SARS-CoV-2 test (green), sample collection for first positive test (pink), transfer to the COVID-19 ward (dark blue) and clinical outcome [death (black), hospital discharge (orange)] for each of the 21 infected patients. The patients numbers are coloured according to the phylogenetic reconstruction (as in Figure 2A), where numbers coloured in grey indicate samples for which virus genome data could not be obtained. To facilitate the identification of patient-to-patient contact, patients timelines (rows in the figure) are grouped by room (identified by capital letters, see Figure S1). The hospitalization days are highlighted in light blue and the room numbers are indicated on the day a given patient was transferred to that room (the room stay extends until a new transfer appears on the timeline). With the exception of patient 35 (Ward A), all patients and room numbers are relative to Ward B. The presence in other hospital areas is also indicated when relevant [Hemodialysis (⊕), Emergency department (‡)]. The date of sample collection for healthcare workers (HCWs) testing positive is also presented at the bottom. The timeline presented starts two weeks before the large test screening and extends until the date of sample collection of the last outbreak-related positive case. Light grey fields (on both sides of the timeline) are included to show events occurring outside the study timeline. †List of HCWs with a positive test on large screening: 1,2,4,5,11,13,14,17,18,22,24,27,28,29,31,33 and 40 (no sequencing data). *, Patient admission before study period; **, Patient admission and last negative test before study period.

### Investigation of nosocomial transmission of SARS-CoV-2 with in-depth contact tracing and viral genomic data

An in-depth investigation was carried out to identify contacts during the 15-day period before the large screening. We collected vast data regarding HCW visits to patient’s rooms, patient’s room transfer and circulation to other treatment areas, which, in general, reflected a complex contact network commonly found in highly “populated” hospital wards. While HCW-patient and patient-patient contacts could be robustly traced back (Figure 1; Figure S1), the frequent contact between HCWs in common areas (inside and outside the ward B) and vast equipment/material sharing led us to assume a high likelihood of contact/transmission between all HCWs in ward B during the evaluated period. To strengthen the investigation, positive RNA samples were sent to the National Institute of Health for SARS-CoV-2 genome sequencing and analysis.

### Genomic characterization of the SARS-CoV-2 outbreak variant

High-quality SARS-CoV-2 genome sequences were acquired from 39 positive samples, enrolling 22 out of the 27 infected HCWs and 17 out of 21 infected patients (Figure 2; Supplementary Table S1). Phylogenetic reconstruction revealed that all cases were part of the same transmission chain, thus confirming a single origin behind this nosocomial outbreak (Figure 2A). The outbreak-associated SARS-CoV-2 variant belongs to COG-UK lineage B.1.1 and Nextstrain clade 20B, carrying the Spike amino acid changes D614G and L176F (Supplementary Table S2). It diverges from the clade 20B root by seven SNPs, five of which being shared with two SARS-CoV-2 viruses (Portugal/PT1550/2020 and Portugal/PT1614/2020) also identified in Lisbon and Tagus Valley region in June-July 2020 (https://insaflu.insa.pt/covid19/; as of 11 December 2020) (Figure 2A; Supplementary Table S2). This suggests that the virus leading to the nosocomial outbreak is a descendent variant that had been circulating in this geographical region for several months. The introduction in the hospital ward B might have occurred a few weeks before the large screening, which is supported by considerable phylogenetic ramification and mutation accumulation during the nosocomial outbreak (five SNPs from the common ancestor to the more divergent virus) (Figure 2A). In fact, the phylogenetic reconstruction revealed two main descendent branches (both with further sub-branch diversification), enrolling nine (cases 24-31 and 39; yellow-like) and 14 (cases 11-23 and 38; blue-like) cases, and six singletons diverging from the ancestral genetic profile (cases 1-10, green) (Figure 2A). Notably, all clusters enrolling identical genetic profiles included both patients and HCWs.

**Figure 2.**
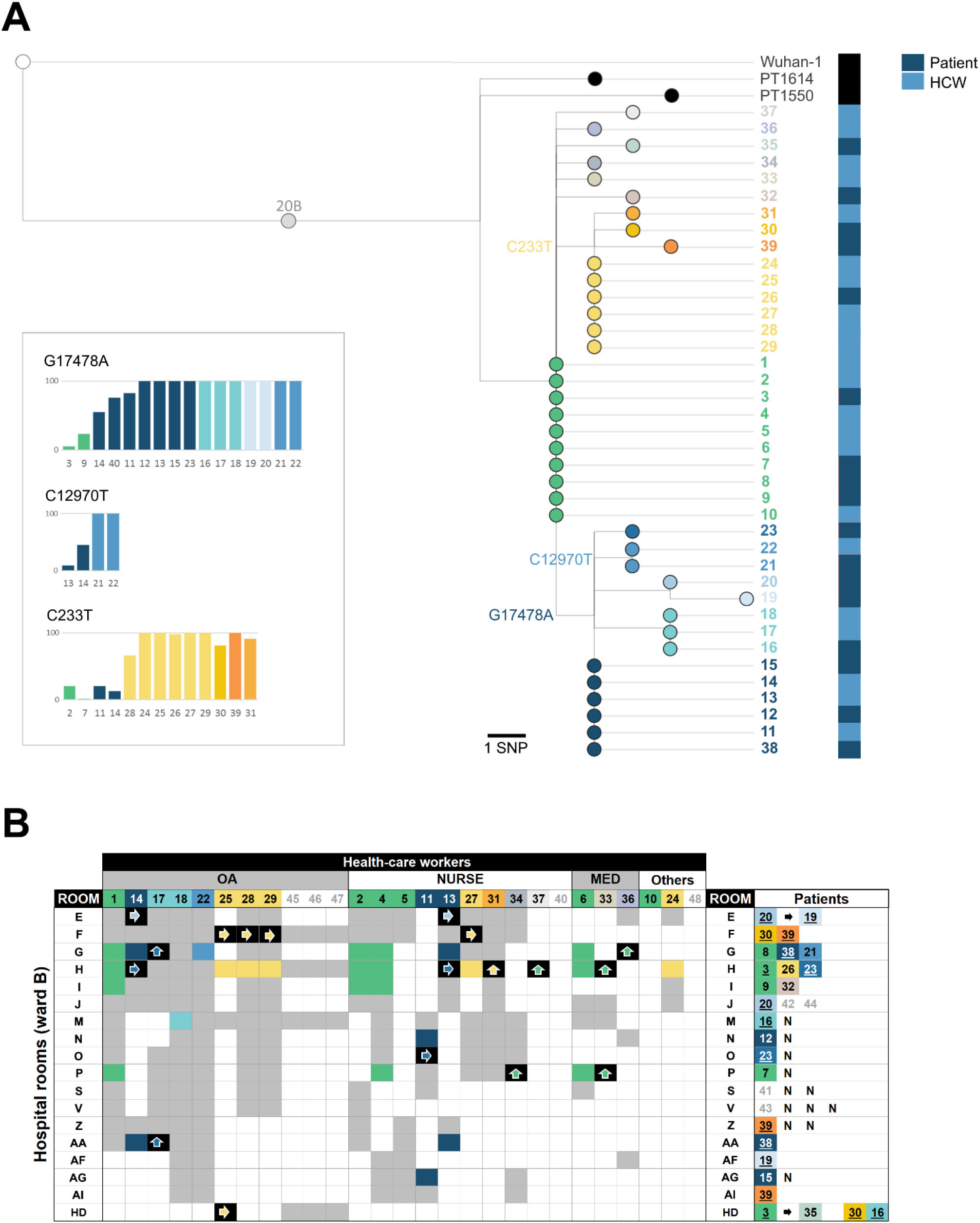
Combined genomic and epidemiological investigation of a SARS-CoV-2 nosocomial outbreak. **A**. Maximum likelihood phylogenetic tree of SARS-CoV-2 genome sequences analysed during the investigation of SARS-CoV-2 positive cases detected in a hospital ward in August 2020, Portugal. The tree includes viral consensus sequences obtained from 22 out of the 27 infected health-care workers (HCWs) and 17 out of 21 infected patients (colored nodes). To provide better phylogenetic context, the two closest sequences of the outbreak SARS-CoV-2 variant detected in Portugal (Portugal/PT1550/2020, collected in 21 June 2020, and Portugal/PT1614/2020, collected in 8 July 2020; https://insaflu.insa.pt/covid19/; as of 9 November 2020) and the SARS-CoV-2 reference genome sequence (used as root; Wuhan-Hu-1/2019; MN908947.3) were also included. Bar graphs next to the tree depict the within-patient frequency fluctuation of three SNP phylogenetic markers (also highlighted in the respective branch) that were detected either as minor variants (<50% in frequency) or as (nearly-)fixed variants in viral populations collected from distinct infected individuals (bars are colored according to the phylogenetic placement of the respective SARS-CoV-2 consensus sequences). The SNP profile of the outbreak-related SARS-CoV-2 is provided in Supplementary Table S2. **B**. HCW-patient contacts occurring during the 15-day period before the large screening. HCW and patient codes are colored according to the phylogenetic placement of the respective SARS-CoV-2 consensus sequences (panel A), with exception of those without SARS-CoV-2 genome data (gray numbers in white fields). The “contact tracing matrix” summarizes the visits of infected HCWs to patient’s rooms (more details about the timeline of the visits is provided in Figure S1) (colored fields). Fields are colored according to virus sequence similarity between HCW and patients, as follows: i) if a given a HCW visited a room with a patient with identical viral consensus sequence (i.e., same genetic profile), the field is colored according to the genetic profile (as in panel A) (high likelihood of direct transmission); ii) if the contact involved ascendent/descendent genetic profiles (plausible transmission) and there was no other contact with an individual with an identical virus genetic profile, the field is colored in black with a colored arrow indicating the potential direction of transmission; iii) if the contact HCW-patient is not supported by viral genetic evidence (implausible transmission), the fields are colored in gray. HD, hemodialysis. N, patients testing negative for SARS-CoV-2; Underlined patient number means that the patient stayed in different rooms during the study period.

### Reconstructing SARS-CoV-2 nosocomial transmission trajectories

The integration of genomic and epidemiological data indicated that SARS-CoV-2 nosocomial dissemination was mainly driven by HCW-patient and HCW-HCW transmission (Figure 2B; Figure S1). Specifically, this scenario is supported by: i) the frequent detection of identical consensus viral sequences among HCWs and between HCW and patients contacting with each other; ii) absence of identical viral sequences in patients that shared the same room; and, iii) detection of identical viral sequences collected from patients that did not contact with each other. Indeed, patient-to-patient direct transmission was more limited, which is also corroborated by the frequent observation of one single patient testing positive in rooms with more than one patient (which occurred in eight rooms) (Figure 2B; Figure S1). Notwithstanding, in a few cases, we found strong genetic evidence of direct patient-to-patient transmission. For instance, besides the “patient 3 - patient 35” transmission during hemodialysis, patient 20 most likely transmitted SARS-CoV-2 to patient 19 during their joint isolation (both were also infected with *Clostridium difficile*) (Figure 2). In order to disclose intricate transmission events, we sought to verify whether mutations accumulating during the transmission chain (phylogenetic markers) were already present as minor variants (sub-populations) in samples with ascendant or identical consensus virus sequences. Remarkably, increasing frequencies were detected in three mutations (C233T, C12970T and G17478A) that turned out to fix throughout the nosocomial outbreak (Figure 2A), For instance, the mutation G17478A (synonymous SNP in ORF1ab / NSP13) defining one of the two main branches (detected at frequencies 55.5-100.0% in that sub-cluster) was already present at a frequency of 5.5% (patient 3) and 23.7% (patient 9) in individuals presenting the viral ancestral profile. This observation suggests, for instance, that patient 3 was most likely one of the first infected individuals in the outbreak and places patient 14 in the potential origin of the “G17478A” sub-cluster transmission chain. Likewise, the mutation C12970T (synonymous SNP in ORF1ab / NSP9), which was found with a minor frequency in HCWs 13 and 14 (9% and 45%, respectively), became fixed (100%) in the HCW 22 and patient 21, likely indicating consecutive direct transmission enrolling these individuals (concordantly, all HCWs 13, 14, and 22 contacted each other and visited patient 21 during the study period). Finally, although the mutation defining the other main sub-branch (C233T; extragenic SNP in 5’ UTR) was already present as a minor variant in the ancestral populations, it was also unexpectedly detected in samples with consensus sequences falling within the other main branch (G17478A). This observation illustrates how difficult it can be to interpret sub-populations transmission and frequency fluctuations when retrospectively reconstructing outbreaks detected after massive screening at a given time point (i.e., the viral population signature at the time of testing may not reflect the population profile at the time of transmission/infection). Even though, our minor variants analysis provided valuable clues about probable transmission trajectories that would be hard to disclose using solely epidemiological and consensus sequence data, especially in a scenario of a complex contact network enrolling a multitude of transmission events.

### Searching for the index case

Considering that most infected individuals were detected by large screening (likely a few weeks after the virus introduction in hospital ward B), it is highly challenging to identify the “index” case of this nosocomial outbreak. According to the timeline (Figure 1), contact tracing data and virus phylogenetic analysis (Figure 2; Figure S1), we can indeed launch several plausible hypotheses. Among the infected patients presenting the most ancestral viral sequences (patients 3, 7, 8 and 9), we hypothesize that patient 3 and 8 have a higher likelihood of being the index case as they were admitted to hospital (testing negative) a few days before patients 7 and 9. In particular, patient 3 should have been infected shortly before or just after the hospital admission, as she/he most likely transmitted SARS-CoV-2 to patient 35 (his/her single contact) during hemodialysis six days later (Figures 1 and 2). Likewise, several HCWs might have also been the index case, namely HCW 1, 2, 4, 5, 6 or 10. If one assumes that the index case is a HCW that transmitted SARS-CoV-2 to a patient with the same viral genetic profile at the beginning of the study period (namely to patient 3 or 8), we can point out HCW 2 and 6 as the most likely HCWs being the index as they visited these patients’ rooms on day 2 and 3 of the study period (Figure 2; Figure S1). In contrast, HCWs 1 and 4 visited those patients on day 5 and HCWs 5 and 10 have not contacted with any patient presenting identical viral sequences (Figure 2). Finally, given the time span between the infection and testing, we cannot completely exclude as a potential index case patients or HCWs showing a virus variant directly descendant from the most ancestral one. In this case, the additional mutations detected at time of testing would represent within-patient evolution after the first transmission events. While this scenario would fit, for instance, the hypothesis of patient 35 (first patient testing positive) being the index case (i.e., patient 35, coming from ward A, would have introduced the virus in ward B by contacting patient 3 during hemodialysis), this is unlikely considering that patient 35 tested negative on day 7 of the study period (Figure 1). Indeed, as stated above, assuming this hypothesis would considerably shrink the outbreak timeline, making it incongruent with the observed virus genetic diversification.

### Outbreak follow-up: implemented measures and impact on subsequent outbreaks

The combined genomic and epidemiological investigation unveiled a scenario of rapid and unnoticed SARS-CoV-2 dissemination within the hospital ward B, mostly driven by HCW-to-HCW or HCW-patient/patient-HCW transmission. In this context, this outbreak led to the implementation of novel measures to minimize the risk of introduction and dissemination of SARS-CoV-2 within a “non-COVID” ward. In particular, communication and educational activities were reinforced to promote compliance with standard precautions, emphasizing the need of frequent hand hygiene, maintenance of safe physical distance (1.5-2 meters) and mandatory use of respiratory protection within the ward (at least a surgical mask). Targeted and specific training events among hospital employees and visitors were carried out and further divulgation was performed through hospital intranet and website, as well as by displaying information at strategic places of the facility. Furthermore, the movement of people within the facility was limited and restricted to essential and staff break periods were recommended to be made individually. The maximum capacity of common areas, namely break rooms, HCWs changing rooms and elevators, was also reduced. Other administrative measures included changing the layout of the hospital cafeteria, with a maximum of two people allowed per table (with a distance of 1.5 meters). All HCWs and visitors were instructed about the need to wear a mask immediately after finishing meals and to avoid staying in these meal places longer than the period required. Visits to all internal medicine wards were canceled during the outbreak period and up to 28 days after the last positive case. As an indirect observation of the success of implementing such measures, subsequent SARS-CoV-2 positive cases within the same non-COVID-19 ward were detected more timely, leading to limited transmission chains (Supplementary Figure S2) In fact, virus genome sequencing of 11 out of 15 new positive cases detected between 9 September and 1 October not only confirmed that the these cases were not related with the previous outbreak, but also that they represented four independent introductions with none or very limited transmission (Supplementary Figure S2).

## CONCLUSION

In the present study, SARS-CoV-2 genome data was key to confirm that a single introduction of SARS-CoV-2 in the non-COVID-19 ward, followed by prolonged silencing transmission, resulted in 48 infected individuals (27 HCW and 21 inpatients). As reported for other nosocomial COVID-19 outbreaks, we observed a high mortality rate (57.1%) among patients, which certainly reflects the comorbidities and demographics of the patients’ population [10, 12]. The combination of vast contact tracing disclosed a complex scenario of SARS-CoV-2 transmission, consolidating the expectation that highly “populated” hospital wards, marked by extensive circulation of HCWs, pose great challenges to reconstruct direct transmission events and the exact virus spread trajectory. These challenges were even more pronounced given: i) the SARS-CoV-2 relatively low substitution rate (around one mutation every 2 weeks) (https://nextstrain.org/ncov/global), which renders identical or nearly-identical sequences in several individuals enrolled in the transmission chain; ii) that most cases were only detected upon massive screening, meaning that the viral genetic profile detected at the time of testing may not reflect the one at the time of transmission/infection; and iii) that most cases were asymptomatic at time testing, hampering any reconstruction based on the chronology of incubation period and symptoms onset. We cannot discard that mild disease symptoms might have occurred (particularly among HCWs), having been disregarded in the context of HCWs high workload and fatigue during COVID-19 crisis, as widely reported [8, 13, 23, 24]. Notwithstanding, our results clearly demonstrated that SARS-CoV-2 dissemination within the hospital ward was mostly mediated by HCW-to-HCW and HCW-patient/patient-HCW transmission. This emphasizes the need for compliance with the current guidelines for infection prevention and control and preparedness for COVID-19 in healthcare settings, given the HCWs increased risk of virus exposure, intensive circulation within hospital facilities and close contact with frail patients [5, 6]. In the studied outbreak, as currently recommended [6], all patients were tested upon admission to hospital and massive laboratory testing of ward residents and staff was performed after the detection of a first case. Nonetheless, the observed scenario of prolonged silencing transmission points that periodic screening of HCWs and patients, even in the absence of known cases, could have anticipated the detection of the nosocomial outbreak. Importantly, the reinforcement of control and prevention measures during and after this outbreak was shown to be effective towards the mitigation of transmission and a more timely detection and containment of novel nosocomial outbreaks. Altogether, the present study shows the benefit of combining genomic and epidemiological data for the investigation of complex nosocomial outbreaks, while providing valuable data to minimize the risk of transmission of COVID-19 in healthcare facilities.

## Data Availability

SARS-CoV-2 genome sequences generated in this study were uploaded to GISAID database (https://www.gisaid.org/).

## Ethical statement

Samples were obtained as part of the routine clinical care procedures of the Hospital Vila Franca de Xira. HCW and patient data were anonymized. This study is covered by the ethical approval issued by the Ethical Committee (“Comissão de Ética para a Saúde”) of the Portuguese National Institute of Health.

## Supplementary Material

**Figure S1.**
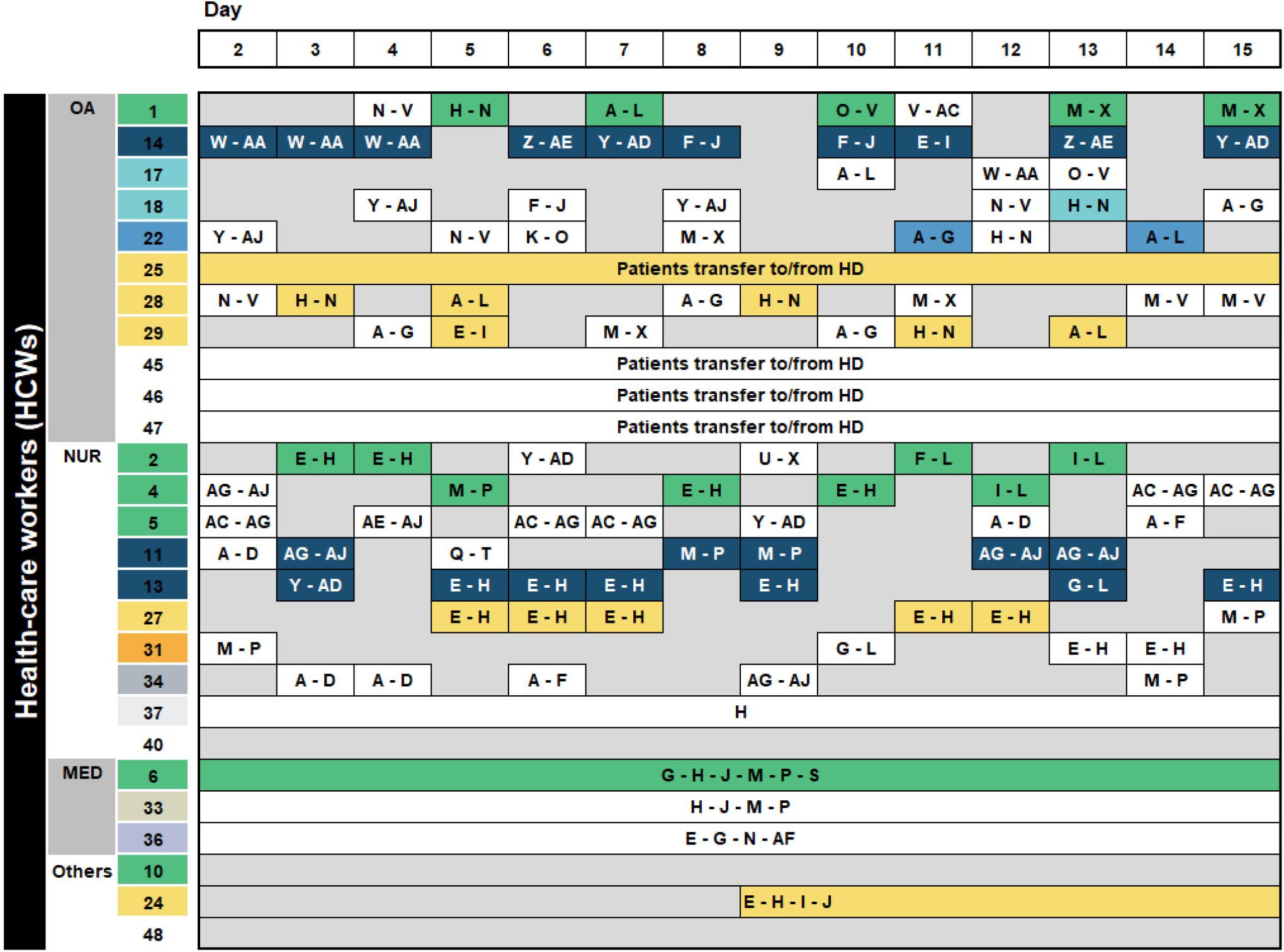
Timeline of HCW-patient contacts before the large screening. HCW and patient codes are colored according to the phylogenetic placement of the respective SARS-CoV-2 consensus sequences (Figure 1A), which exception of those without SARS-CoV-2 genome data (gray numbers in white fields). The timeline (matching that of Figure 1, in which day 1 corresponds to the first day of the 15-day period before the large screening) summarizes the visits of infected HCWs to patient’s rooms (identified by capital letters, in alphabetical order, from A to Z, followed by AA to AJ). Whenever a HCW visited a room with a patient with identical viral consensus sequence (i.e., same genetic profile), the field is colored according to the genetic profile (as in Figure 1A) (i.e., cases with the highest likelihood of direct transmission). HD, hemodialysis.

**Figure S2.**
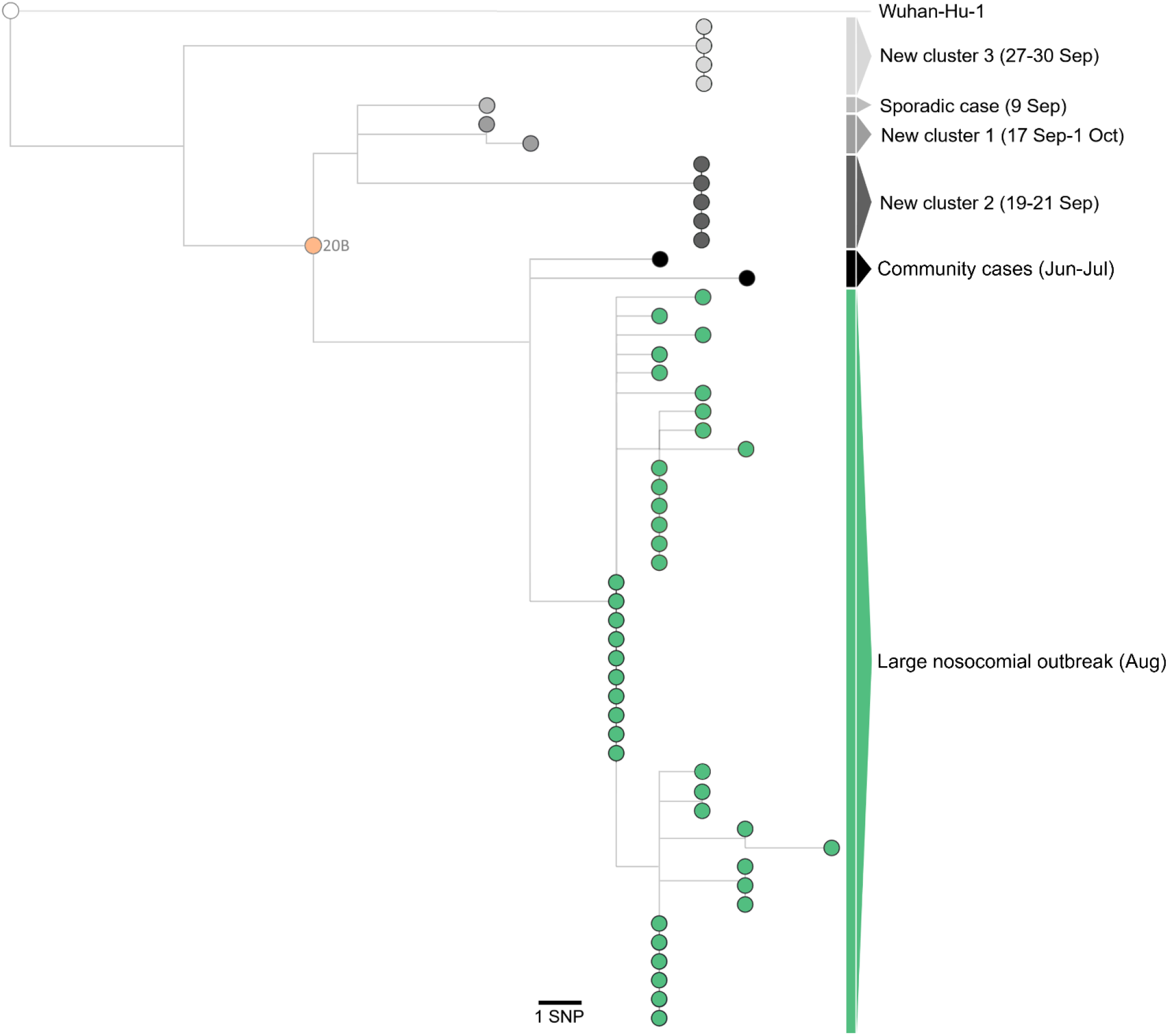
Independent hospital introductions of SARS-CoV-2 cases after the large nosocomial outbreak led to limited transmission chains. Maximum likelihood phylogenetic tree of SARS-CoV-2 genome sequences analysed during the investigation of SARS-CoV-2 positive cases detected in a non-COVID-19 ward of a large hospital in Portugal, between August and October 2020. The tree includes viral consensus sequences analyzed during the large nosocomial outbreak (n = 40) in August (presented in the same order as in Figure 2), as well as 11 out of 15 new positive cases detected between 9 September and 1 October. To provide better phylogenetic context, the two closest sequences of the outbreak SARS-CoV-2 variant detected in community (Portugal/PT1550/2020, collected in 21 June 2020, and Portugal/PT1614/2020, collected in 8 July 2020; https://insaflu.insa.pt/covid19/; as of 9 November 2020) and the SARS-CoV-2 reference genome sequence (used as root; Wuhan-Hu-1/2019; MN908947.3) were also included.

**Supplementary Table S1.** List of outbreak-related SARS-CoV-2 genome sequences generated in this study.

| Sample designation | Patient / HCW code | GISAIID Accession ID |
| --- | --- | --- |
| Portugal/PT1701/2020 | 1 | EPI_ISL_693630 |
| Portugal/PT1702/2020 | 2 | EPI_ISL_693631 |
| Portugal/PT1703/2020 | 3 | EPI_ISL_693632 |
| Portugal/PT1704/2020 | 4 | EPI_ISL_693633 |
| Portugal/PT1705/2020 | 5 | EPI_ISL_693634 |
| Portugal/PT1706/2020 | 6 | EPI_ISL_693635 |
| Portugal/PT1707/2020 | 7 | EPI_ISL_693523 |
| Portugal/PT1708/2020 | 8 | EPI_ISL_693636 |
| Portugal/PT1709/2020 | 9 | EPI_ISL_693520 |
| Portugal/PT1710/2020 | 10 | EPI_ISL_693637 |
| Portugal/PT1711/2020 | 11 | EPI_ISL_693638 |
| Portugal/PT1712/2020 | 12 | EPI_ISL_693639 |
| Portugal/PT1713/2020 | 13 | EPI_ISL_693640 |
| Portugal/PT1714/2020 | 14 | EPI_ISL_693641 |
| Portugal/PT1715/2020 | 15 | EPI_ISL_693642 |
| Portugal/PT1716/2020 | 16 | EPI_ISL_693643 |
| Portugal/PT1717/2020 | 17 | EPI_ISL_693644 |
| Portugal/PT1718/2020 | 18 | EPI_ISL_693645 |
| Portugal/PT1719/2020 | 19 | EPI_ISL_693646 |
| Portugal/PT1720/2020 | 20 | EPI_ISL_693647 |
| Portugal/PT1721/2020 | 21 | EPI_ISL_693648 |
| Portugal/PT1722/2020 | 22 | EPI_ISL_693649 |
| Portugal/PT1723/2020 | 23 | EPI_ISL_693519 |
| Portugal/PT1724/2020 | 24 | EPI_ISL_693650 |
| Portugal/PT1725/2020 | 25 | EPI_ISL_693651 |
| Portugal/PT1726/2020 | 26 | EPI_ISL_693652 |
| Portugal/PT1727/2020 | 27 | EPI_ISL_693653 |
| Portugal/PT1728/2020 | 28 | EPI_ISL_693654 |
| Portugal/PT1729/2020 | 29 | EPI_ISL_693518 |
| Portugal/PT1730/2020 | 30 | EPI_ISL_693524 |
| Portugal/PT1731/2020 | 31 | EPI_ISL_693655 |
| Portugal/PT1732/2020 | 32 | EPI_ISL_693537 |
| Portugal/PT1733/2020 | 33 | EPI_ISL_693656 |
| Portugal/PT1734/2020 | 34 | EPI_ISL_693539 |
| Portugal/PT1735/2020 | 35 | EPI_ISL_693534 |
| Portugal/PT1736/2020 | 36 | EPI_ISL_693657 |
| Portugal/PT1737/2020 | 37 | EPI_ISL_693536 |
| Portugal/PT1738/2020 | 38 | EPI_ISL_693516 |
| Portugal/PT1739/2020 | 39 | EPI_ISL_693535 |

**Supplementary Table S2.** Genome background of the outbreak-related SARS-CoV-2.

| Genome position<sup>a</sup> | Nucleotide change | Amino acid change |
| --- | --- | --- |
| **241<sup>b,c</sup>** | C>T | (5'UTR) |
| **1457<sup>c</sup>** | C>T | NSP2: R218C (non-syn) / ORF1a: R398C (non-syn) |
| **3037<sup>b,c</sup>** | C>T | NSP3: F106F (syn) / ORF1a: F924F (syn) |
| **6500** | C>T | NSP3: P1261S (non-syn) / ORF1a: P2079S (non-syn) |
| **7006<sup>c</sup>** | C>T | NSP3: T1429T (syn) / ORF1a: T2247T (syn) |
| **14408<sup>b,c</sup>** | C>T | NSP12b: P314L (non-syn) / ORF1b: P314L (non-syn) |
| **22088<sup>c</sup>** | C>T | S: L176F (non-syn) |
| **23403<sup>b,c</sup>** | A>G | S: D614G (non-syn) |
| **24019<sup>c</sup>** | A>G | S: E819E (syn) |
| **28846<sup>c</sup>** | C>T | N: R191R (syn) |
| **28881-3<sup>b,c</sup>** | GGG>AAC | N: R203K, G204R (non-syn) |
| **29692** | G>T | (3'UTR) |

<sup>a</sup> Genome positions refer to the reference SARS-CoV-2 Wuhan-Hu-1/2019 sequence (GenBank accession MN908947).
<sup>b</sup> Nextstrain 20B clade SNP markers.
<sup>c</sup> Mutations shared with not-related community-circulating virus (Portugal/PT1550/2020 and Portugal/PT1614/2020) detected in Lisbon and Tagus Valley region in June-July 2020 (https://insaflu.insa.pt/covid19/; as of 11 December 2020)
syn: synonymous mutation.
non-syn: non-synonymous mutation.

## References

1. Wu F, Zhao S, Yu B, et al. A new coronavirus associated with human respiratory disease in China. Nature 2020; 580(7803):E7. https://doi.org/10.1038/s41586-020-2202-3

2. Wang X, Zhou Q, He Y, et al. Nosocomial outbreak of COVID-19 pneumonia in Wuhan, China. Eur Respir J. 2020; 55(6):2000544. doi: https://doi.org/10.1183/13993003.00544-2020

3. World Health Organization (WHO) Coronavirus disease (COVID-19) Weekly Epidemiological Update – 08 December 2020. https://www.who.int/emergencies/diseases/novel-coronavirus-2019/situation-reports/.

4. Hu B, Guo H, Zhou P, Shi ZL. Characteristics of SARS-CoV-2 and COVID-19. Nat Rev Microbiol 2020; 6:1– 14. doi: https://doi.org/10.1038/s41579-020-00459-7

5. World Health Organization (WHO). Infection prevention and control during health care when coronavirus disease (COVID-19) is suspected or confirmed. Geneva: World Health Organization; 2020 (available at: https://apps.who.int/iris/rest/bitstreams/1284718/retrieve).

6. European Centre for Disease Prevention and Control. Infection prevention and control and preparedness for COVID-19 in healthcare settings – Fifth update. 6 October 2020. ECDC: Stockholm; 2020. https://www.ecdc.europa.eu/sites/default/files/documents/Infection-prevention-and-control-in-healthcare-settings-COVID-19_5th_update.pdf

7. The Lancet. COVID-19: protecting health-care workers. Lancet 2020; 395(10228):922. https://doi.org/10.1016/S0140-6736(20)30644-9

8. Black JRM, Bailey C, Przewrocka J, et al. COVID-19: the case for health-care worker screening to prevent hospital transmission. Lancet 2020; 395(10234):1418–1420. https://doi.org/10.1016/S0140-6736(20)30917-X.

9. Zhan M, Qin Y, Xue X, Zhu S. Death from Covid-19 of 23 Health Care Workers in China. N Engl J Med 2020; 382(23):2267–2268. https://doi.org/10.1056/NEJMc2005696

10. Lucey M, Macori G, Mullane N, et al. Whole-genome sequencing to track SARS-CoV-2 transmission in nosocomial outbreaks. Clin Infect Dis 2020; ciaa1433. https://doi.org/10.1093/cid/ciaa1433

11. Heinzerling A, Stuckey MJ, Scheuer T, Xu K, Perkins KM, Resseger H, Magill S, Verani JR, Jain S, Acosta M, Epson E. Transmission of COVID-19 to Health Care Personnel During Exposures to a Hospitalized Patient - Solano County, California, February 2020. MMWR Morb Mortal Wkly Rep 2020; 69(15):472–476. https://doi.org/10.15585/mmwr.mm6915e5

12. Rickman HM, Rampling T, Shaw K, Martinez-Garcia G, Hail L, Coen P, Shahmanesh M, Shin GY, Nastouli E, Houlihan CF. Nosocomial transmission of COVID-19: a retrospective study of 66 hospital-acquired cases in a London teaching hospital. Clin Infect Dis. 2020; ciaa816. https://doi.org/10.1093/cid/ciaa816

13. Rivett L, Sridhar S, Sparkes D, et al. Screening of healthcare workers for SARS-CoV-2 highlights the role of asymptomatic carriage in COVID-19 transmission. Elife 2020; 9:e58728. https://doi.org/10.7554/eLife.58728

14. Meredith LW, Hamilton WL, Warne B, et al. Rapid implementation of SARS-CoV-2 sequencing to investigate cases of health-care associated COVID-19: a prospective genomic surveillance study. Lancet Infect Dis 2020; 20(11):1263–1272. https://doi.org/10.1016/S1473-3099(20)30562-4

15. Safdar N, Moreno GK, Braun KM, Friedrich TC, O’Connor DH. Using Virus Sequencing to Determine Source of SARS-CoV-2 Transmission for Healthcare Worker. Emerg Infect Dis 2020; 26(10):2489–2491. https://doi.org/10.3201/eid2610.202322

16. Sikkema RS, Pas SD, Nieuwenhuijse DF, et al. COVID-19 in health-care workers in three hospitals in the south of the Netherlands: a cross-sectional study. Lancet Infect Dis 2020; 20(11):1273–1280. https://doi.org/10.1016/S1473-3099(20)30527-2

17. Quick J, Grubaugh ND, Pullan ST, et al. Multiplex PCR method for MinION and Illumina sequencing of Zika and other virus genomes directly from clinical samples. Nat Protoc 2017; 12(6):1261–1276. https://doi.org/10.1038/nprot.2017.066

18. Borges V, Isidro J, Cortes-Martins H, et al. Massive dissemination of a SARS-CoV-2 Spike Y839 variant in Portugal. Emerg Microbes Infect 2020; 2:1–58. doi: https://doi.org/10.1080/22221751.2020.1844552

19. Borges V, Pinheiro M, Pechirra P, Guiomar R, Gomes JP. INSaFLU: an automated open web-based bioinformatics suite “from-reads” for influenza whole-genome-sequencing-based surveillance. Genome Med 2018; 10(1):46. https://doi.org/10.1186/s13073-018-0555-0

20. Hadfield J, Megill C, Bell SM, et al. Nextstrain: real-time tracking of pathogen evolution. Bioinformatics 2018; 34(23):4121–4123. https://doi.org/10.1093/bioinformatics/bty407

21. Mercatelli D, Triboli L, Fornasari E, Ray F, Giorgi FM. Coronapp: A web application to annotate and monitor SARS-CoV-2 mutations. J Med Virol 2020; https://doi.org/10.1002/jmv.26678

22. Rambaut A, Holmes EC, O’Toole Á, et al. A dynamic nomenclature proposal for SARS-CoV-2 lineages to assist genomic epidemiology. Nat Microbiol 2020; 5(11):1403–1407. https://doi.org/10.1038/s41564-020-0770-

23. Alharbi J, Jackson D, Usher K. The potential for COVID-19 to contribute to compassion fatigue in critical care nurses. J Clin Nurs 2020; 29(15-16):2762–2764. https://doi.org/10.1111/jocn.15314

24. Shanafelt T, Ripp J, Trockel M. Understanding and Addressing Sources of Anxiety Among Health Care Professionals During the COVID-19 Pandemic. JAMA 2020; 323(21):2133–2134. https://doi.org/10.1001/jama.2020.5893

